# Alcohol related consequences and disparate outcomes in college students at a Hispanic Serving Institution

**DOI:** 10.1101/2022.12.20.22283742

**Authors:** Wayne Kepner, Guadalupe X. Ayala, James Lange, John Elder

## Abstract

**Objective:** Examine the relationship between alcohol consumption and alcohol related consequences (ARCs) for college students at a Hispanic Serving Institution (HSI).

**Participants:** Random sample of 375 students at a large university in the pacific south-west.

**Methods:** Cross sectional study using an online version of the ACHA-NCHA II survey. Chi-square tests and stepped logistic regression models were conducted to explore the relationship between race/ethnicity and the experience of any alcohol-related consequence, adjusting for covariates.

**Results:** Among heavy drinkers, Hispanic students, compared to White students, were 65.9% less likely to report one or more ARC (OR= 0.34, 95%CI = 0.15, 0.79).

**Conclusions:** Given the severity of alcohol related consequences in the college population, the potential protective environmental effects of attending a HSI warrant further study.

## Introduction

Young adults in the college community are at an increased risk for experiencing an alcohol-related consequence (ARC) including alcohol abuse and alcohol use disorder (AUD) (Carter et al., 2010). Broadly speaking, ARCs encompass three major domains: (1) damage to self, (2) damage to others, (3) institutional damage (Perkins, 2002). Among U.S. college students, ARCs account for a large proportion of unintended injury and death (Shillington & Clapp 2001), including motor-vehicle crashes. Approximately 1,248 college students aged 18-24 die each year from alcohol-related injuries (Hingson et al., 2005). Additionally, every year 696,000 students are assaulted by another student who has been drinking, and 97,000 students report experiencing alcohol-related sexual assault or date rape (White & Hingson, 2013).

Ethnic minorities make up a large proportion of the overall college population in the U.S., but they also face a disproportionate amount of disparities in health-related incidence rates of many chronic illnesses such as diabetes, certain cancers and heart disease (Keppel, 2007).Unique elements of campus environment such as the availability of alcohol, the presence of Greek communities, and even the percentage of minority students have been shown to influence student drinking habits (Lui et al., 2020). In the college population, Straka and colleagues have shown an association between social exclusion and a desire for “belonging” in minority groups with alcohol use (Straka, Gaither, Acheson, & Swartzwelder, 2019). Furthermore, in a study of 5,369 students on two college campuses, LaBrie et al., (2012) demonstrated a possible interaction between the size of the minority population on campus and drinking levels of ethnically diverse students. Des Rosiers et al. demonstrated that assimilated (i.e. those who adopt U.S. practices and customs) Hispanic students reported engaging in higher levels of risky alcohol behaviors (e.g. binge drinking and unprotected sex) compared to Hispanic students who retained more of their cultural heritage (Des Rosiers et al., 2013).

Consequently, there are inequitable outcomes between racial/ethnic minorities and White students(Paschall & Flewelling, 2002). Wagner and colleagues demonstrated in 2002 that Black students were more likely to report a persistent desire to cut down on their alcohol consumption as well as unsuccessful attempts to quit alcohol compared to White students. Similarly, U.S. born Hispanic students were more likely to report continued drinking despite persistent social or legal troubles (Wagner et al., 2002). A study conducted by Welte & Barnes in 1987 found that American Indians are highest in “per capita alcohol consumption, percentage of heavy drinkers, number of times drunk, number of alcohol-related problems and illicit drug use.” Welte & Barnes also found that in general, minority groups experience more problems than Whites when holding constant the amount of alcohol that they consume. Although research shows that AAPI generally consume less alcohol, and also have fewer ARCs compared to White students, there is great heterogeneity in the prevalence rates of alcohol consumption in the APPI population (Luczak et al., 2006) which makes such generalizations problematic; similar problems exist in the characterizations of Hispanic populations (Elder et al., 2009). However, Nguyen et al. found that at the same level of drinking, AAPI students experienced greater levels of incapacitated rape compared to their White student counterparts (Nguyen et al., 2010). Asian students have also been shown to experience greater rates of alcohol-related blackouts, but this association was not supported for White, Hispanic or Black students (Gonçalves et al., 2017).

A study by (Blume, Lovato, Thyken, & Denny, 2012) showed that ethnic minority students in predominantly white universities are at a greater risk of certain alcohol consequences. This study looked at the associations between microaggressions, self-efficacy, binging-drinking events and alcohol consequences--as defined by the Rutgers Alcohol Problems Index. Blume, Lovato, Thyken & Denny showed that ethnic minority students in predominantly white universities experience a greater number of micro-aggressions, which they found was significantly associated with an increased level of anxiety and a greater number of alcohol problems. Likewise, Barry et al., (2017) reported that black men attending a predominantly white institution (PWI) reported a significantly higher level of alcohol consumption along with significantly more mental health conditions. Furthermore, the same study reported that attendance at a minority-serving institution was associated with fewer reports of mental health conditions among black male students. Studies have shown that Latinx students attending PWIs have different drinking patterns compared to Latinx students who attend Hispanic serving institutions (HSI) (Vaughan et al., 2015). Specifically, Vaughan and colleagues reported that the perceptions of other’s drinking were more strongly linked to Latinx students personal drinking at non-Hispanic serving institutions. Consequently, the researchers posit that protective effect provided by HSI may come in the form of a culturally affirming environment.

Regarding alcohol consumption and alcohol related consequences, the college campus and environment are unique elements of society. The current literature is inconsistent regarding the influence of race/ethnicity and ARCs. While there are trends that show for example, that Black and Hispanic students are less likely to report high levels of drinking and yet face similar rates of ARC’s, there is evidence to suggest that race/ethnicity is not significantly associated with ARCs.

### Present study

In this study, we aimed to explore the association between race/ethnicity and ARCs at a HSI. We hypothesized that minority students (i.e. Non-White Hispanic, Non-White/Non-Hispanic) would benefit from the socially diverse environment with a reduction in the number of endorsed ARCs. Conversely, we hypothesized that the majority population (i.e. Non-Hispanic Whites) would not benefit from a similar reduction in ARCs compared to national averages. Consequently, our research question is as follows: Does race/ethnicity moderate the relationship between alcohol consumption and alcohol related consequences (ARCs) for college students at a Hispanic Serving Institution (HSI).

## Methods

### Study Design

This cross-sectional study is a secondary data analysis of the 2018 American College Health Association’s National College Health Assessment II (ACHA-NCHA II). In 2015, the California State University (CSU) Chancellor’s office informed all CSU campuses that the ACHA NCHA II would be implemented bi-annually throughout all CSU campuses. This survey was deployed by a large public university in the Pacific Southwest for the first time in 2016, and again in 2018.

### Data Source

Since 1920, the American College Health Association (ACHA) has served as a lead organization and advocacy group for the advancement of college and university health (American College of Health Association, 2005). ACHA provides education, communications, and products and services, as well as promotes research and culturally competent practices, to enhance its members’ ability to advance the health of all students and campus communities (American College Health Association, 2018). ACHA represents over 10 million college students in 1,100 institutions of higher education and boasts a diverse membership with representation from two- and four-year schools, public and private, small and large and minority serving institutions.

The ACHA-NCHA is a survey that was created in 2000 to assist college health service providers, counselors, health educators and administrators to evaluate students’ habits, behaviors and perceptions on several physical and mental health domains (American College Health Association, 2018). Researchers, health professionals and student administrators use the ACHA-NCHA data to design and evaluate health promotion programs, allocate staff and fiscal resources for campus health education programs, provide data for campus task forces, create grant proposals, etc. (Rahn et al., 2016). Institutions self-select to administer the survey either on an annual or bi-annual survey.

ACHA-NCHA was pilot tested in 1998-1999 and evaluated for reliability and validity through comparisons to national studies such as the National College Health Risk Behavior Survey (Douglas et al., 1997), the Harvard School of Public Health College Alcohol Study (Wechsler & Nelson, 2008), and the U.S. Department of Justice: The National College Women Sexual Victimization Study 2000 (American College Health Association, 2004; 2005; Rahn et al., 2016). The ACHA collects NCHA data and provides detailed analysis along with executive summaries to participating institutions (American College Health Association, 2016; American College Health Association, 2017). These analyses include student demographics, safety and violence statistics, depression and suicide data, adverse childhood experiences, reports of use and abuse of alcohol, tobacco, and other drugs, sexual behaviors, Body Mass Index and nutrition, physical activity, access to health information, and health status. Furthermore, the ACHA aggregates the data of all participating institutions into larger datasets called “reference groups” which may be used for secondary analyses through a formal approval process overseen by ACHA (American College Health Association, 2018). However, since participating members are self-selected, the data may not generalize to all schools in the United States (Research Projects and Data Access, n.d.). The ACHA-NCHA can be taken either online via Qualtrics or through a paper-only version.

In 2008, the ACHA created an updated version of the survey called the ACHA-NCHA II which includes emerging health issues such as tobacco use with a water pipe, un-prescribed prescription drug use and new birth control products (American College Health Association, 2008). Also, questions were deleted which were deemed irrelevant such as student credit card use patterns; response options were also amended (e.g. “select all that apply”). Initially, these revisions were meant as minor updates, but the extent of the changes led to the creation of a second version of the NCHA. The revisions were informed by data collected from the first 8 years of the survey (i.e., 2000 to 2008). To date, the ACHA-NCHA II is the largest known nationwide dataset pertaining to college students’ health (American College Health Association, 2008).

### Sample recruitment and eligibility criteria

The data used for this study were collected from the web-based version of the ACHA-NCHA II administered to college students in the Spring of 2018 from March 14^th^ to April 8^th^, 2018. The campus Registrar’s office randomly selected 7,000 SDSU undergraduate students who were then contacted by ACHA via email with an invitation link to take the online version of the ACHA-NCHA II. In the same email, students were informed about a lottery incentive which included one grand prize of a $150 gift card to Starbucks and thirteen other prizes to the campus bookstore (1-$100, 4-$50 and 8-$25 cards). Odds of winning a prize were estimated in the email as being 1 in 50, and students who received the invitation but who did not wish to complete the survey were also eligible to enter the lottery by emailing the sponsoring department – the Well Being and Health Promotion Department. Students were informed that their responses would be completely de-identified and confidential, and that any personal data collected through this process such as email addresses would be deleted by ACHA at the completion of their survey. Students were also encouraged to both skip items that they were uncomfortable answering or to omit sensitive data which they felt might identify them. Finally, the email included a written consent portion which explained the sensitive nature of some of the items on the ACHA-NCHA II and provided students with information about how to contact the Counseling and Psychological Center if they wished to discuss any issues raised by the survey. The initial survey email was followed up every few days with a reminder email to the students over the course of approximately 3 weeks. In total, from among the 7,000 students who received the email, 665 students completed the survey which resulted in a response rate of 9.5%.

## Measures

The ACHA-NCHA II is a 66-item web-based survey. The following topics are included in the ACHA-NCHA II: alcohol, tobacco, and other drug use; sexual health; weight, nutrition, and exercise; injury prevention; personal safety and violence; physical and psychological health (American College Health Association, 2018). The items are asked using a variety of response formats such as Likert-scales, list selection, and yes/no responses. The time frame covered by items range from lifetime prevalence to “within the last 2 weeks” (American College Health Association, 2018). The purpose of this study was to assess the association of race/ethnicity on ARCs through the analysis of self-reported survey data from college students who reported consumption of alcohol.

### Alcohol consumption

For our exposure variable “alcohol consumption,” we used a survey item that asked participants, “The last time you “partied”/socialized, how many alcoholic drinks did you have?”. Participants were able to indicate an exact number of drinks consumed. Invalid responses were those with either no response or multiple responses, and they were removed from analysis by the ACHA. Researchers who utilize ACHA-NCHA data often prefer to report the quantity of drinks consumed “the last time they partied” which also informed our coding (Barry et al. 2017; Moore et al., 2013; Siebert et al., 2003). The alcohol quantity responses were used to divide the respondents into three separate categories: abstainers, low to moderate drinkers and heavy drinkers. The three alcohol consumption categories were created using drinking guidelines which define low-risk drinking and high-risk drinking. Specifically, the low to moderate drinkers category was created using the U.S. Department of Health and Human Services drinking guidelines which defines low-risk drinking as one drink per day for women and two drinks per day for men (U.S. Department of Health and Human Services, 2015). The National Institute on Alcohol Abuse and Alcoholism (NIAAA) sets the standard for moderate risk drinking as up to four alcoholic beverages for males and three alcoholic drinks for females in any single day.Consequently, female students who reported between one and three drinks, and male participants who reported between one and four drinks the last time they partied were coded as “low to moderate drinking.” For the heavy drinking category, the Substance Abuse and Mental Health Services Administration (SAMHSA) states that consuming more than four drinks in one occasion for females and five drinks in one occasion for males is associated with a higher risk of negative consequences such as legal problems and physical withdrawal symptoms (SAMHSA, 2019). Consequently, female participants who reported over four or more drinks in one occasion, and male participants who reported five or more drinks per occasion were coded as “heavy drinkers.”

### Alcohol related consequences

To assess the consequences of drinking, nine items were presented in the ACHA-NCHA II. All nine questions began with the stem: “Within the last 12 months, have you experienced any of the following when drinking alcohol.” The nine options were: (1) “Did something you later regretted?”, (2) “Forgot where you were or what you did?”, (3) “Physically injured yourself?”, (4) “Seriously considered suicide?” (5) Got in trouble with the police?”, (6) Someone had sex with me without my consent?”, (7) Had sex with someone without their consent?”, (8) Had unprotected sex?”, (9) Physically injured another person?”. All nine questions had three response options: 1 = “N/A, Don’t Drink, 2 = “No”, and 3 = “Yes”. Since this study is concerned with alcohol related consequences, eligible responses were limited to “Yes” or “No”. Those who endorsed “N/A, Don’t Drink” were coded as “abstainers” and were excluded from our analysis (n=14). An ARC scale was then created for each eligible respondent which represented the sum score of consequences. Finally, respondents were categorized into one of two groups (0) “No Alcohol Related Consequences”, and (1) “Any Alcohol Related Consequences”. The Cronbach’s alpha for this ARC scale was 0.659.

### Race/Ethnicity

For our independent variable, “Race/Ethnicity”, we used a survey item that asked participants, “How do you usually describe yourself?” The response options were 1 = “White”, 2 = “Black or African American”, 3 = “Hispanic or Latino/a”, 4 = “Asian or Pacific Islander”, 5 = “American Indian or Alaskan”, 6 = “Biracial or Multi Racial”, and 7 = “Other”. For this analysis, race/ethnicity was divided into three categories: 1 = “White”, 2 = “Hispanic”, and 3 = “Non-White, Non-Hispanic.” A subset of respondents indicated that they were both “White” and “Hispanic,” these students were coded as “White.” Participants who did not report race or ethnicity were excluded from analysis (n=2).

### Covariates

Age, gender, alcohol consumption and Greek affiliation have been identified as predictors of alcohol related consequences during a student’s college years (Borsari, Murphy, & Barnett, 2007; Dennhardt & Murphy, 2013). For example, research shows that first-year male students engage in higher levels of heavy-risk drinking compared to females and older students (Borsari et al., 2007). Male college students are also more likely to experience aggression, justice involvement, and property destruction compared to college females (Borsari et al., 2007).

Research also suggests that initiation of drinking under the age of 21 is associated with experiencing alcohol related consequences (Hingson et al., 2000), and that delaying the initiation of drinking for 5 years can reduce the negative consequences of drinking by upwards of 50% (Dawson et al., 2008). Consequently, we excluded students whose self-reported age was greater than 24 (n=147) to focus on a specific period of social development—namely young adulthood in the college environment. Research shows that students who report low to moderate amounts of alcohol consumption have lower risks of being pushed, hit or assaulted or experiencing an unwanted sexual advance compared to those who engage in high levels of alcohol consumption (Mellins et al., 2017). In terms of Greek affiliation, students in fraternities and sororities experience greater alcohol related harms than non-Greek affiliated students (Barry, 2007).

## Data analyses

SAS 9.4 was used to generate descriptive statistics and chi-square tests to explore differences between race/ethnicity, age, gender, Greek affiliation, and alcohol consumption for students who reported experiencing any ARCs compared to students who did not experience any ARCs. A stepped logistic regression model was then conducted to explore the relationship between race/ethnicity and the experience of any alcohol-related consequence, adjusting for covariates. The chi-square tests and stepped logistic regression models were used to explore the main research question: Does race/ethnicity moderate the relationship between alcohol consumption and alcohol related consequences for college students at a Hispanic Serving Institution? Step one included all covariates (age, gender, alcohol consumption, Greek affiliation), step two included covariates and the race/ethnicity variable, and step three included all covariates, races/ethnicities, and a race/ethnicity x alcohol consumption interaction term. For our regression models we reported odds ratios (ORs) with 95% confidence intervals (CIs). Potential covariates were included in a multivariable model based on previous research indicating that age, gender, alcohol consumption, and Greek affiliation are associated with ARCs. All covariates (age, gender, alcohol consumption, Greek affiliation) were assessed for multicollinearity, but none were found to be highly correlated with each other (Pearson’s r<0.3).

## Results

### Descriptive statistics

The final analytical sample consisted of 375 participants. Table 1 presents demographic information about study participants, including differences between participants who reported at least one ARC compared to those who reported no ARCs. Within this sample, 78.7% of students were female and the mean age was 20.66(SD=1.72) years. For this sample, 20.8% of students identified as Hispanic, 22.1% identified as Non-White/Non-Hispanic (which included Asian, Black, American Indian, Alaskan Native, Native Hawaiian, Bi-racial or Other), and 57.1% identified as Non-Hispanic White.

**Table 1.**
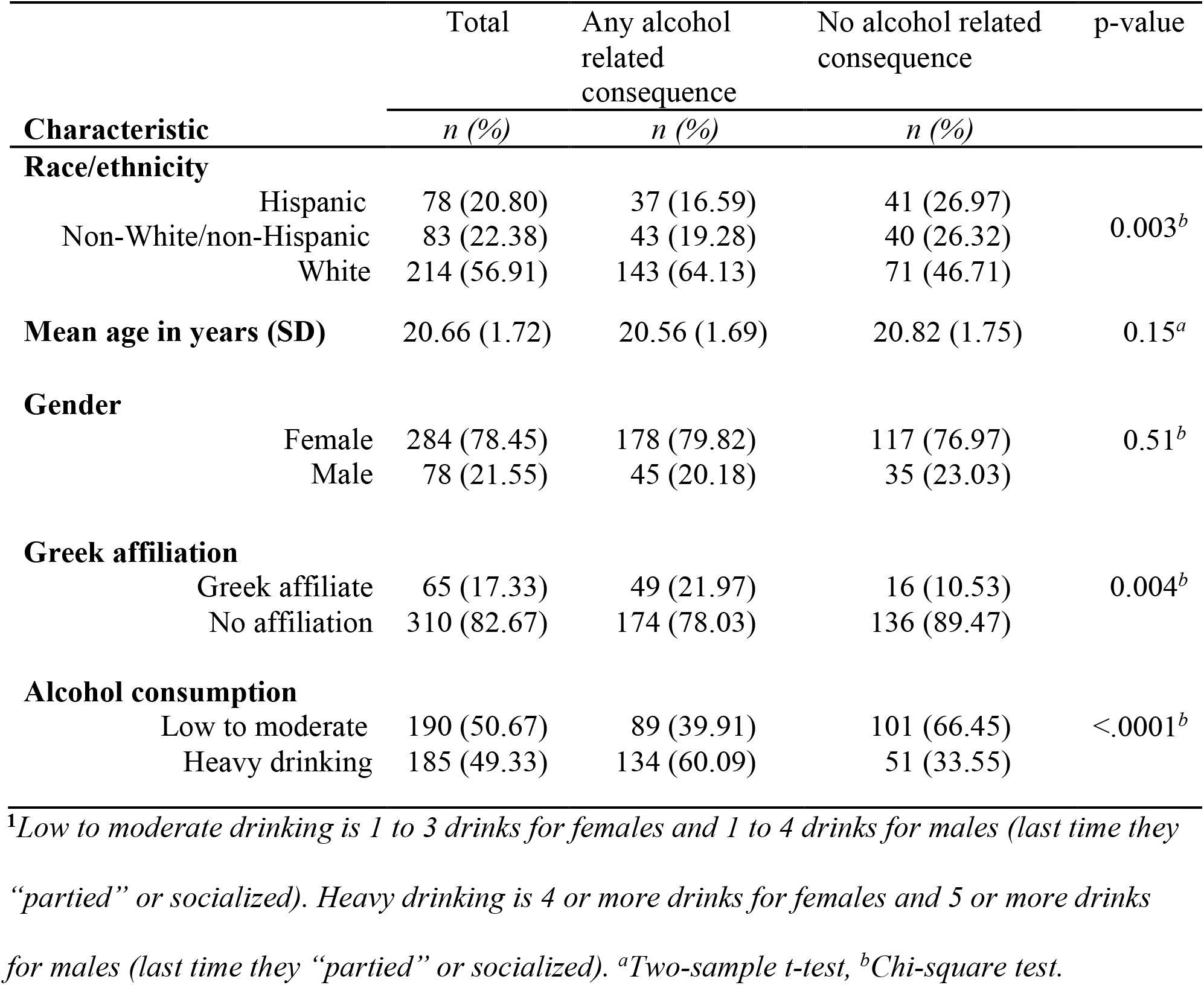
Demographic characteristics of student drinkers in the spring of 2018 at a HSI presented by the ARC variable (n=375).

|  |  | Total | Any alcohol<br>related<br>consequence | No alcohol related<br>consequence | p-value |
| --- | --- | --- | --- | --- | --- |
| Characteristic |  | *n (%)* | *n (%)* | *n (%)* |  |
| **Race/ethnicity** |  |  |  |  |  |
|  | Hispanic | 78 (20.80) | 37 (16.59) | 41 (26.97) | 0.003<sup>b</sup> |
|  | Non-White/non-Hispanic | 83 (22.38) | 43 (19.28) | 40 (26.32) |  |
|  | White | 214 (56.91) | 143 (64.13) | 71 (46.71) |  |
| **Mean age in years (SD)** |  | 20.66 (1.72) | 20.56 (1.69) | 20.82 (1.75) | 0.15<sup>a</sup> |
| **Gender** |  |  |  |  |  |
|  | Female | 284 (78.45) | 178 (79.82) | 117 (76.97) | 0.51<sup>b</sup> |
|  | Male | 78 (21.55) | 45 (20.18) | 35 (23.03) |  |
| **Greek affiliation** |  |  |  |  |  |
|  | Greek affiliate | 65 (17.33) | 49 (21.97) | 16 (10.53) | 0.004<sup>b</sup> |
|  | No affiliation | 310 (82.67) | 174 (78.03) | 136 (89.47) |  |
| **Alcohol consumption** |  |  |  |  |  |
|  | Low to moderate | 190 (50.67) | 89 (39.91) | 101 (66.45) | <.0001<sup>b</sup> |
|  | Heavy drinking | 185 (49.33) | 134 (60.09) | 51 (33.55) |  |

<sup>1</sup>Low to moderate drinking is 1 to 3 drinks for females and 1 to 4 drinks for males (last time they “partied” or socialized). Heavy drinking is 4 or more drinks for females and 5 or more drinks for males (last time they “partied” or socialized). <sup>a</sup>Two-sample t-test, <sup>b</sup>Chi-square test.

There were no age or gender difference in those who experienced ARCs compared to those who did not experience ARCs (both n.s.). Among students who self-reported as Hispanic, 50% experienced ARCs and 50% did not experience any ARCs. Among those who self-reported as White, 69.1% experienced ARCs and 30.9% did not experience any ARCs. Among students who self-reported as Non-White/Non-Hispanic, 46.9% experienced ARCs and 53.1% did not experience any ARCs. In terms of differences by Greek affiliation, significantly more students who experienced ARCs reported belonging to a fraternity or sorority (21.9 %) than students who did not experience ARCs (10.5%, p=0.004). In addition, significantly more students who experienced ARCs reported engaging in heavy drinking (60.1%) compared to those who did not experience ARCs (33.6%, p <0.001).

### Distribution of alcohol related consequences

The mean ARC score was 1.43 (SD=1.53), and the range was six with an interquartile range of two. As shown in Table 2, more than half of students (59.01%) reported either zero (38.2%) or one ARC (20.8%). Only 5.8% of students experienced either five (3.1%) or six (1.7%) ARCs. Approximately 88.1% of students experienced less than four ARCs total as a result of their alcohol consumption.

**Table 2.** Distribution of the summed scores from the alcohol related consequences scale (n=375)

| **Alcohol related**<br>**consequence score** | **Frequency** | **Percent** |
| --- | --- | --- |
| 0 | 138 | 38.23 |
| 1 | 75 | 20.78 |
| 2 | 67 | 18.56 |
| 3 | 38 | 10.53 |
| 4 | 26 | 7.20 |
| 5 | 11 | 3.05 |
| 6 | 6 | 1.66 |

### Stepped logistic regression

As shown in Table 3, step one included known covariates (age, gender, alcohol consumption, and Greek affiliation), step two included the covariates along with race/ethnicity, and step three included the covariates, race/ethnicity, and an interaction term between race/ethnicity and alcohol consumption (i.e. low to moderate drinker or heavy drinker).

**Table 3.** Multivariate stepped logistic regression with dichotomous ARC as dependent variable; step one, two and three.

| Variable | Step One |  | Step Two |  | Step Three |  |
| --- | --- | --- | --- | --- | --- | --- |
|  | aOR | 95% CI | aOR | 95% CI | aOR | 95% CI |
| **Age** | 0.93 | 0.82 - 1.05 | 0.94 | 0.82 – 1.06 | 0.939 | 0.827 – 1.07 |
| **Gender** |  |  |  |  |  |  |
| Female | 1.40 | 0.83 - 2.38 | 1.40 | 0.822 – 2.38 | 1.388 | 0.812 – 2.37 |
|  | Ref |  | Ref |  | Ref |  |
| Male |  |  |  |  |  |  |
| **Greek Status** |  |  |  |  |  |  |
| No affiliation | 0.52* | 0.28 - 0.98 | 0.60 | 0.31 – 1.15 | 0.591 | 0.306 – 1.15 |
| Greek affiliate | Ref |  | Ref |  | Ref |  |
| **Number of Drinks<sup>1</sup>** |  |  |  |  |  |  |
| Low to moderate | 0.35*** | 0.23 - 0.55 | 0.60** | 0.24 – 0.58 | 0.278 | 0.151 - 0.510 |
| Heavy drinking | Ref |  | Ref |  | Ref |  |
| **Race/Ethnicity<sup>2</sup>** |  |  |  |  |  |  |
| Hispanic |  |  | 0.551 | 0.32 – 0.96 | 0.784 | 0.382 – 1.61 |
| NWNH |  |  | 0.712 | 0.41 – 1.24 | 0.863 | 0.425 – 1.75 |
|  |  |  | Ref |  | Ref |  |
| White |  |  |  |  |  |  |
| **Alcohol consumption<sup>1</sup> x Race/Ethnicity<sup>2</sup>** |  |  |  |  |  |  |
| *Low to moderate* |  |  |  |  |  |  |
| Hispanic |  |  |  |  | 0.784 | 0.382 – 1.61 |
| NWNH |  |  |  |  | 0.863 | 0.425 – 1.75 |
|  |  |  |  |  | Ref |  |
| White |  |  |  |  |  |  |
| *Heavy drinking* |  |  |  |  | 0.341 | 0.147 – 0.792 |
| Hispanic |  |  |  |  | 0.558 | 0.237 – 1.314 |
| NWNH |  |  |  |  | Ref |  |
| White |  |  |  |  |  |  |
| **R<sup>2</sup>** | 0.11 |  | 0.13 |  | 0.14 |  |

\*p<0.05, \*\*p<0.01, \*\*\*p<0.0001. <sup>1</sup>Low to moderate drinking is 1 to 3 drinks for females and 1 to 4 drinks for males (last time they “partied” or socialized). Heavy drinking is 4 or more drinks for females and 5 or more drinks for males (last time they “partied” or socialized). <sup>2</sup>NWNH = Non-White/non-Hispanic.

For the first step, Greek status (p<0.01) and number of drinks consumed (p≤.001) were significantly associated with experiencing ARCs; age and gender were not. Participants with no Greek affiliation were 48% less likely to experience ARCs (OR = 0.52, 95% CI 0.28 - 0.98). Low to moderate drinkers were 65% less likely to experience ARCs (OR = 0.35, 95% CI 0.23 - 0.55). For the second step, alcohol consumption was the only variable significantly associated with experiencing an ARC (p<0.001). Students who engaged in low to moderate drinking were 40% less likely to experience an ARC compared to those who engaged in heavy drinking (OR=0.60, 95% CI 0.24 – 0.60). For the third step, which included all covariates, race/ethnicity and a race/ethnicity* alcohol consumption interaction term, age, gender, and Greek affiliation were not significant. Among heavy drinkers, Hispanics compared to Whites were 65.9% less likely to report one or more ARCs (OR= 0.34, 95%CI = 0.15, 0.79). All other comparisons for race/ethnicity * alcohol consumption interactions were non-significant and the maximum likelihood test for the overall interaction term was non-significant. This means that there was no significant interaction between race/ethnicity and alcohol consumption with respect to the experience of ARCs for White students, Hispanic students and Non-White/Non-Hispanic students among low to moderate drinkers. For heavy drinkers, there was no significant difference between White students and Non-White/Non-Hispanic students and similarly no significant difference between Non-White/Non-Hispanic students and Hispanic students.

## Discussion

The present study builds upon previous research on alcohol related disparities in the college population. Presently, there are only a few studies that have focused specifically on ethnic/racial factors of ARCs at HSIs. This study intended to fill this research gap by addressing the question: Does race/ethnicity moderate the relationship between alcohol consumption and alcohol related consequences (ARCs) for college students at a Hispanic Serving Institution (HSI)? In this study, participants who engage in heavy drinking and participants with Greek affiliation are more likely to experience ARCs. Among Hispanic, Non-Hispanic/Non-White, and White participants, Hispanic participants were less likely to experience ARCs. After adjusting for age, sex, alcohol consumption and Greek affiliation, a significant interaction effect was demonstrated between race/ethnicity and alcohol consumption. Specifically, among heavy drinkers, Hispanic students were less likely to report ARCs compared to White students. This interaction suggests that Hispanic students who engage in heavy drinking are less likely to experience an ARC compared to White students who are also heavy drinkers.

### Comments

Contrary to the results from our study, previous research suggest that Hispanic students experience more problems because of heavy drinking when compared to non-Hispanic Whites. Mulia and colleagues demonstrated in 2009 that African American and Hispanic drinkers were significantly more likely than white drinkers to report consequences of drinking even after adjusting for differences in demographic characteristics (Mulia et al., 2009). Most strikingly, the racial/ethnic disparities reported in that study were most prominent among the individuals reporting little or no heavy drinking; this disparity disappeared at the highest levels of heavy drinking. Hispanic participants were three times as likely to report ARCs compared to White participants at low levels of drinking. In our study, there was no discrepancy between races/ethnicities at low to moderate levels of drinking, but instead there was a significant interaction in which Hispanic participants who engaged in heavy drinking experienced less consequences than White participants at similar levels of heavy drinking. Mulia and colleagues explained that racial/ethnic stigma and poverty were the two most relevant factors for the experience of ARCs among their participants. Since our study sampled a relatively heterogenous population attending school at a diverse institution, the effects of racial/ethnic stigma and poverty may have had less impact.

In a study by Vaughan et al., 2015, the results indicated a significant interaction between attending an HSI and the perception of the number of drinks consumed by a “typical student.” For students attending a non-Hispanic serving institution, the perception of other students’ drinking behavior more strongly predicted personal drinking. Early research in the field of college alcohol consumption has shown that the social environment can significantly affect the drinking patterns of individuals; gender composition of the social group and the number of individuals in the group are important components of this social dynamic (Aitken, 1985; Carman, 1977). Regarding alcohol consequences and decision-making, research suggests that group dynamics and pro-social roles (Lange, Devos-Comby, Moore, Daniel, & Homer, 2011) within drinking groups can affect alcohol consumption and outcomes. Consequently, Vaughan and colleagues suggested that the protective effect of attending an HSI may be related to the existence of a more culturally affirming college atmosphere. In 2008, Corbin, Vaughan & Fromme showed that peer influence was an important factor for both Latino students and Caucasian students with respect to drinking patterns. However, the same study found that family influences were also significant in Latinx youths-particularly Latina women (Corbin, Vaughan & Fromme, 2008). These associations between social context and drinking behavior is a possible explanation for the reduced experience of ARCs by Hispanic heavy drinkers. The pro-social influence of family on Hispanic students may partially explain our results.

Our study expands on the topic of racial/ethnic disparities by suggesting that Hispanic race/ethnicity is associated with a modest decrease in likelihood of experiencing ARCs for heavy drinkers at a HSI. A negative association approaching significance was demonstrated between Hispanic race/ethnicity and experiencing ARCs, and a significant interaction effect was demonstrated between heavy drinking behavior and race/ethnicity (OR= 0.34, 95%CI = 0.15, 0.79. Although our study did not assess social norms, acculturation, or family environment, the implications of these studies may partially explain our findings. African Americans who have faced discrimination are 50% more likely to smoke and report heavier alcohol use (Borrell et al., 2010). Similarly, Hispanics reported reporting racial/ethnic discrimination were reported to have 60% greater odds of heavy drinking. In 2015, Cheng & Mallinckrodt demonstrated that Hispanic students at HSI’s face lower levels of racial/ethnic discrimination compared to Hispanic students at primarily white institutions. In 2012, Labrie and colleagues demonstrated that the size of a minority population relative to other students moderated the relationship between drinking and social norms which may also be true of ARCs. Consequently, unidentified factors such as a culturally affirming environment may have a role in lowering the odds of experiencing ARCs in Hispanic students who engage in heavy drinking at a HSI.

### Strengths and limitations

One of the major strengths of this study is the ability of researchers, administrators, staff, and faculty to apply these findings directly to the population from which the data was collected. The ability to attribute national-level findings directly to a specific population has limitations depending on study design. The variation in campus size, culture and demographic make-up are limitations for perfect generalizability to individual environments. Consequently, for programs, grants and policies at the organizational level, organizational specific data are appropriate and may hold more utility for community member and local stakeholders.

Many limitations in this survey exist that limit generalizability to other institutions. Primarily, the ACHA is not a probability sample because many institutions which implement the ACHA survey are self-selecting (although non-members can also choose to participate); this limits generalizability. Furthermore, the over-representation of Caucasian and female students in this sample does not match the enrollment population at this HSI. This limitation is especially important considering that one of the main points of exploration for this study was Race/Ethnicity. Also, since gender is a moderator of alcohol consequences, the over representation of females makes this analysis limited in its generalizability. Another limitation is the low response rate of 9.5% for the overall data collection period which still managed to result in a total sample of over 650 students. The low response rate may partially explain the lack of representation found in the previously mentioned demographic groups. Also, a low response rate may indicate that not all students were comfortable answering questions for the school which may represent a self-selection bias, or a healthy-worker effect for the respondents that did enroll in the study on top of the existing social desirability bias. Regarding timeframes, this survey is limited in that it asks for consequences over the last 12 months. There is a potential for a recall bias in this way since a year is a long time to remember. Also, since college students also drink alcohol at home and on breaks, when using a 12-month time-frame there is less assurance that these consequences are actually a result of being on or near campus. Possible covariates of interest that were not considered include financial status and type of resident. Finally, this sample of students from a HSI in southern California may not be generalizable to students at other HSIs across the country. For example. the within-group variability of Hispanic students including generation status, immigration status, place of birth or country of origin were not accounted for in our analysis.

Furthermore, a limitation exists related to the wording of the consequence items. In our survey, “doing something that you later regretted” can have different interpretations. Student’s interpretations of consequences can vary which also has a direct impact on their behavior. Studies have shown that the perceived positivity of ARCs are associated with higher levels of alcohol consumption, blackouts and regretted sex (Mallet et al., 2008). Mallet and colleagues demonstrated that approximately 25% of students evaluated a hangover as positive which was also true of 12% of students who had experienced a blackout. Our studies ARC counts may be higher than reported if students chose not to endorse the engaging in “regrettable behavior” item after experiencing a hangover. Any cultural diversions in item response behavior for particularly stigmatized, traumatic, or shameful incidents also plays an important factor in our analysis which we were unable to account for.

Likewise, there may be an attribution error occurring with the students’ responses due to the phrasing of the question. For example, the question asked, “As a result of your drinking, have you experienced the following consequences.” However, a student may not be able to accurately attribute their drinking to that negative consequence. In other words, they may not be able to discern whether it was the drinking that made them fall, the wet sidewalk, or maybe it was purely an accident. For a deeper analysis of this topic, one would require more pertinent information than is provided from this survey. For example, the students’ socio-economic status would be helpful and may be a potential confounder for this study. Also, geographic location and social makeup of the drinking scenario or naturalized drinking group (NDG) is also an important facet which was not covered in this study (Lange et al., 2011).

Finally, there are numerous explanations as to the reason why Hispanic participants reported less ARCs which may be explained by measurement error. For example, Hispanic heavy drinkers in our sample might be overrepresented in certain school organizations such as athletic teams which are risk factors for ARCs (Safer & Piane, 2007). Hispanic students could be less likely to live on-campus which would mean they are less likely to be monitored by campus authorities and thus less likely to experience ARCs. Socio-economic factors such as parental income was also unaccounted for in this analysis. Errors in drink measurements have been also been demonstrated in which students underestimated the amount of alcohol consumed leading to ethnic/racial differences in number of drinks reported (Kerr & Greenfield, 2007).

### Implications

If a culturally affirming atmosphere is protective against ARC’s for Hispanic heavy drinkers, the precise mechanism for how students of color prevent higher levels of ARCs may be studied and, if proven effective and appropriate, may be emulated to different campuses or student groups. Further research can focus on what types of consequences are more likely to be faced by heavy drinkers at HSIs, focusing on minority students. The perception, or reality, of getting into trouble at higher levels of intoxication for students of color may preclude them from engaging in behavior which may result in confrontations with police while drinking. Research has shown that Hispanics are more likely than whites to be arrested for drunk driving despite similar rates of driving while drunk (Caetano & Clark, 1998). Further research can also focus on whether racial/ethnic groups employ useful protective behavioral strategies (PBS) against real or perceived consequences of drinking. Studies have shown that less frequent use of PBS (e.g. refraining from drinking games) was associated with increased alcohol related problems (Martens et al., 2004).

## Conclusion

In order to reduce disparities surrounding alcohol consumption in the general population, it may be useful to determine whether certain environments, such as HSI’s, offer any protective benefit to persons of color in order to study that phenomenon and potentially replicate it or foster its growth. Although evidence suggests that Hispanic college students experience more ARC’s than white students, our study demonstrated that among heavy drinkers, Hispanic students are less likely to experience ARCs compared to White students.

## Data Availability

All data produced in the present study are available upon reasonable request to the authors

## Conflict of interest disclosure

The authors report no possible conflicts of interest and no financial relationships with commercial interests.

## Notes

### Competing Interest Statement

The authors have declared no competing interest.

### Funding Statement

This study did not receive any funding

### Author Declarations

IRB of San Diego State University gave ethical approval for this work

